# Trends in Dental Care Visits Across the Lifespan Before and During the COVID-19 Pandemic in the United States: Implications for Practice and Policy

**DOI:** 10.64898/2026.02.07.26345812

**Authors:** Preeti Pushpalata Zanwar, Hossein Zare, Kopal Mathur, Lyubov Slashcheva, Bei Wu

## Abstract

**Objective:** Age-specific disparities in dental care use continue to exist in the United States. The COVID-19 pandemic led to widespread delays in non-urgent dental care. This study provides national estimates of dental care use and examines associated factors for dental visits in the United States population before and during the COVID-19 pandemic.

**Methods & Data Source:** We used the nationally representative Medical Expenditure Panel Survey for the pre-COVID-19 years (2018-2019) and the COVID-19 years (2020-2021). We estimated yearly survey-weighted trends in the mean number of non-zero dental visits by age using an adapted Anderson Behavior and Dental Care Use conceptual model and Poisson regression, controlling for a comprehensive set of confounders across five domains of influence. Dental care visits were defined as visits to any dental care provider.

**Results:** The overall analytic sample included non-institutionalized, community-living persons (unweighted n=6518, weighted N∼320 million), grouped by age as 0-17, 18-44, 45-64, 65-74, and 75+, present in all four years. The prevalence ratio (PR) for dental visits was slightly higher for ages 75+ than for ages 65-74 across 2018-2021, increasing from 1.73 (95% CI: 1.4, 2.1) to 1.84 (95% CI: 1.5, 2.3) to 2.13 (95% CI: 1.7, 2.7) from 2018 to 2020, then rebounding to near pre-pandemic levels in 2021 to 1.66 (95% CI, 1.3, 2.0). Associated factors during the COVID-19 pandemic years 2020-2021 that increased dental visits included dental insurance, high income, and having a usual source of care (p<0.01).

**Conclusions:** Dental care use among older adults increased in 2021; however, it did not fully return to pre-pandemic levels.

**Clinical Implications:** Understanding disparities in associated factors for dental care use across all age groups is critical to improving dental practice, policy, and the provision of timely, high-quality care. Future research should investigate factors contributing to delays in dental care beyond the COVID-19 pandemic, including caries burden, decayed teeth, and tooth loss, particularly among older adults.

## 1. Introduction

The American Dental Association (ADA) recommends regular dental care as an important part of overall health [1,2]. Oral health affects people of all ages. Routine dental visits help prevent oral diseases and cancer, preserve natural teeth, and lower the risk of chronic diseases [3,4,5]. However, not everyone has equal access to dental care, which affects individuals throughout their lives [6,7,8,9]. Parental income and education levels significantly influence children’s access to dental care [8]. Children from low-income families are less likely to receive preventive dental care, which increases their risk of cavities and tooth decay [9]. Many children and adolescents lack dental insurance coverage, despite programs such as Medicaid, the Children’s Health Insurance Program, and the Affordable Care Act [8,9]. Teens may also be more likely to use tobacco or nicotine products and to neglect oral hygiene practices [8]. These behaviors contribute to an increased incidence of tooth decay, gum disease, and orthodontic complications [8,9].

Dental benefits for adults have not improved as much [2]. Working-age adults who lack dental insurance or have limited coverage may delay or avoid dental visits because of cost [10,11]. This can lead to dental problems that worsen over time. Preventive dental care helps adults avoid serious issues such as tooth loss or infections [5,6,7]. Adults with chronic health conditions often require more complex dental care and counseling [12]. For example, people with diabetes are more likely to develop gum disease, which can impact their overall health if left untreated [13,14]. Compared to adults, older adults are more likely to experience complex oral health issues, such as tooth loss, periodontal disease, dry mouth, and oral cancer [15–16]. Additionally, older adults may have underlying health conditions, such as arthritis, dementia, or osteoporosis, that complicate their treatment outcomes [16]. Receiving timely dental care is as important for older adults as receiving medical care.

Dental visits help maintain oral health, support proper nutrition, aid in clear speech, and promote social well-being. Good dental care can also prevent malnutrition and overall decline in health [13,14,15,16,17]. Older adults often face financial and non-financial barriers to accessing dental care. For example, Medicare usually does not cover dental care. Some older adults also have cognitive limitations that make it difficult to seek care [8,18]. Because of these challenges, older adults may have more unmet dental needs or may avoid visiting the dentist [19]. COVID-19 caused financial hardship for both older adults and working-age adults. Many working-age adults’ lost jobs, income, and dental insurance [20]. The pandemic also disrupted access to dental care. Dental offices closed temporarily, and some providers retired. Staff shortages and fears of infection were common. As a result, many people could not access dental care when needed. Nearly half of adults reported having to delay preventive dental care, and many postponed or skipped non-urgent dental visits. During this period, adults with chronic health problems or complex care needs required more dental visits [12,19,20,21,22,23,24,25,26]. Prior studies have used the Medical Expenditure Panel Survey (MEPS) and other data to examine trends in dental visits among the United States (U.S). non-institutionalized civilian population. For example, two reports using MEPS data found that 37%, or nearly 121 million Americans, had at least one dental or medical visit in 2019. However, 18 million fewer Americans used dental care in 2020 than in 2019 [22,26]. Additionally, an ADA report using MEPS data found that 43% of Americans visited a dentist in 2021 [21, 22, 26]. Fifty percent of seniors and children did so, while only 39% of adults ages 19-64 saw a dentist [21,22,26]. Two studies using data on dental practices serving vulnerable Americans from Federally Qualified Health Centers found that although dental care use fully rebounded by August 2020, it remained 8% lower than pre-pandemic levels [23,25]. Dental care also rebounded more slowly than medical care, driven primarily by non-preventive oral care and higher demand for oral surgery and tele-dental care [23,25]. While previous studies examined dental care use by demographics over multiple years of nationally representative surveys [33], few studies have examined a comprehensive set of associated factors and their impacts on dental care use trends before and during COVID-19 across all age groups, including children, adults, middle-aged adults, and older adults.

This study examines the impact of the COVID-19 pandemic on access to dental care among the community-living, civilian, non-institutionalized U.S. population during a period of significant economic disruption. Using four years of Medical Expenditure Panel Survey (MEPS) data, we measured and compared patterns of dental care use for two years before and two years during the onset of COVID-19 (2018-2021). The analysis provides national estimates of trends in mean annual dental care visits and examines associated factors influencing these trends across age groups. We hypothesized that although dental visits would vary by age group, there would be a universal decline during the COVID-19 pandemic. Furthermore, we anticipated that the absence of a complete rebound to pre-COVID levels would indicate persistent structural inequalities in dental access, rather than a temporary response to economic disruption.

## 2. Methods

This research adheres to the Strengthening the Reporting of Observational Studies in Epidemiology (STROBE) Statement for observational study reporting. The Institutional Review Board at Thomas Jefferson University determined the study was exempt from review. As the primary data source, we used the Medical Expenditure Panel Survey (MEPS), a nationally representative dataset widely recognized for its comprehensive information on healthcare utilization in the U.S. population. The methodology and data collection processes of MEPS have been thoroughly described in prior publications [30,31,32,33,34,35].

### 2.1 Study Design

We analyzed data from Panel 23 of the MEPS Household Component, covering 2018 through 2021 [34]. Traditionally, MEPS surveys involve five rounds of in-person interviews with participating households over two years. However, due to the COVID-19 pandemic, data collection for Panel 23 shifted to nine rounds of telephone interviews [35].

### 2.2 Participants and Study Sample

The MEPS-HC 236 file is a de-identified, public dataset that includes individuals and families residing in the community who remained in the U.S. throughout all four study years. Those who were born, died, or left the country for military service during the period were excluded from the analytic sample [30,31,32,33]. Panel 23 initially included 7,080 participants with no age restrictions. We used the YEARDIND==1 variable to identify and retain only those present for the full four-year period, yielding an unweighted sample of 6,518 and a weighted sample of approximately 320 million individuals, representing 92.1% of the original records.

### 2.3 Outcome Measures

Following the method described by Brown et al. (1999), we defined dental service use as the proportion of the population who accessed any dental services during the study period [36]. Dental care utilization was estimated using the DVTOTYEAR variable, which counts office visits to all types of dental professionals, including general dentists, dental hygienists, specialists, dental technicians, dental surgeons, orthodontists, endodontists, and periodontists. For each year, we calculated the mean number of dental visits (excluding those with zero visits) by age group [34,36].

### 2.4 Primary Independent Variable

Recognizing the importance of dental care throughout the life course, we included participants from all age groups, regardless of their dental status [1,2,3,4,5,6,7,8,9,10,11,12,13,14,15,16,17,18,28]. The AGELAST variable provided each person’s age at last eligibility within the study window. Age was grouped into five categories: 0-17.9 years, 18-44.9 years, 45-64.9 years, 65-74.9 years, and 75 years or older. For analytic purposes, the years 2018-2019 were defined as pre-COVID-19, and 2020-2021 as the COVID-19 period.

### 2.5 Covariates

The Andersen Health Behavior Model was adapted to select variables that may affect dental service utilization, grouping them into five domains **(Figure 1)** [37,38,39,40]. These domains were: (1) demographic and social factors (such as age, gender, race/ethnicity, marital status, geographic region, and nativity); (2) healthcare and dental access factors (including dental and health insurance status, educational attainment, and household income); (3) health status indicators (such as cognitive limitations and complete tooth loss); (4) need factors, which included the presence and number of chronic conditions (e.g., diabetes, hypertension, cardiovascular disease, stroke) and the number of prescription medications used; and (5) personal health behaviors, including smoking frequency and whether a person had a usual source of care.

**Figure 1:**
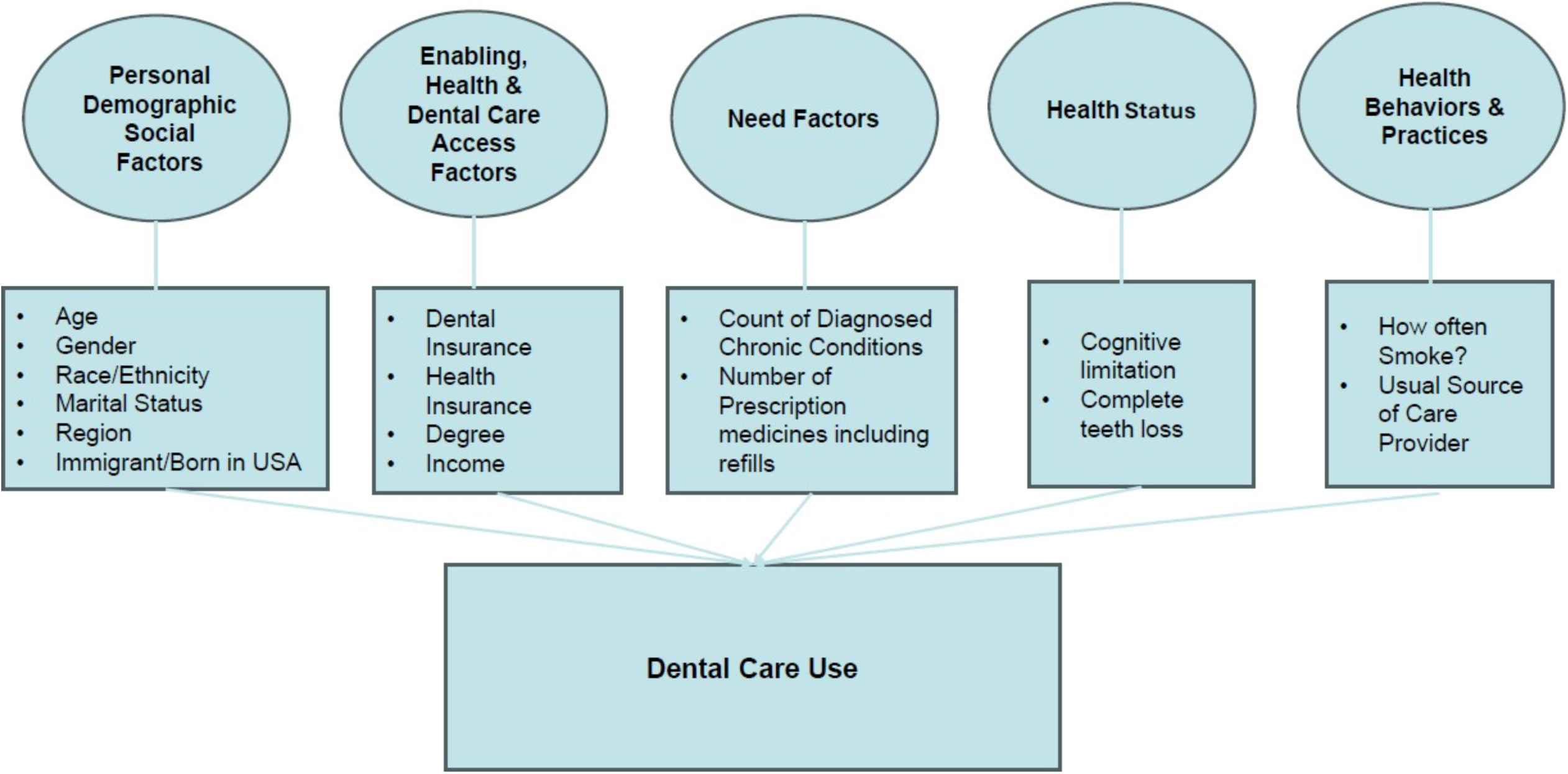
Adapted Anderson Health Behavior Conceptual Framework for Dental Care Use

### 2.6 Statistical Analyses

For each year from 2018 to 2021, we calculated the proportion of individuals in each age group who had zero dental visits and those with at least one visit; age-group differences were evaluated with F-tests. We next estimated the mean number of dental visits (excluding those with zero) by age group and reported 95% confidence intervals for each estimate. Poisson regression models were employed for each year to assess dental care use, first by age and then adjusting for all covariates to identify independent predictors. We also calculated the percent change in the average number of dental visits for each age group over the study period. All analyses accounted for the MEPS complex survey design by applying weights, variance strata, and primary sampling units; results are survey-weighted, and statistical significance was defined as p < 0.01 [32,34]. Analyses were conducted using Stata version 16 [41].

## 3. Results

The analytic sample included 6,518 individuals-children, adolescents, and adults who participated in all four years of the study (2018-2021), representing an estimated 320 million people in the U.S. population aged 0 to 75+ (**Table 1**). Approximately 20%-23% of the sample were children (0-17.9 years), one-third were adults aged 18-44.9 years, over one-quarter were 45–64.9 years, and 15%-17% were 65 years or older. Dental insurance coverage ranged from 43% to 45% during the study period.

**Table 1:**
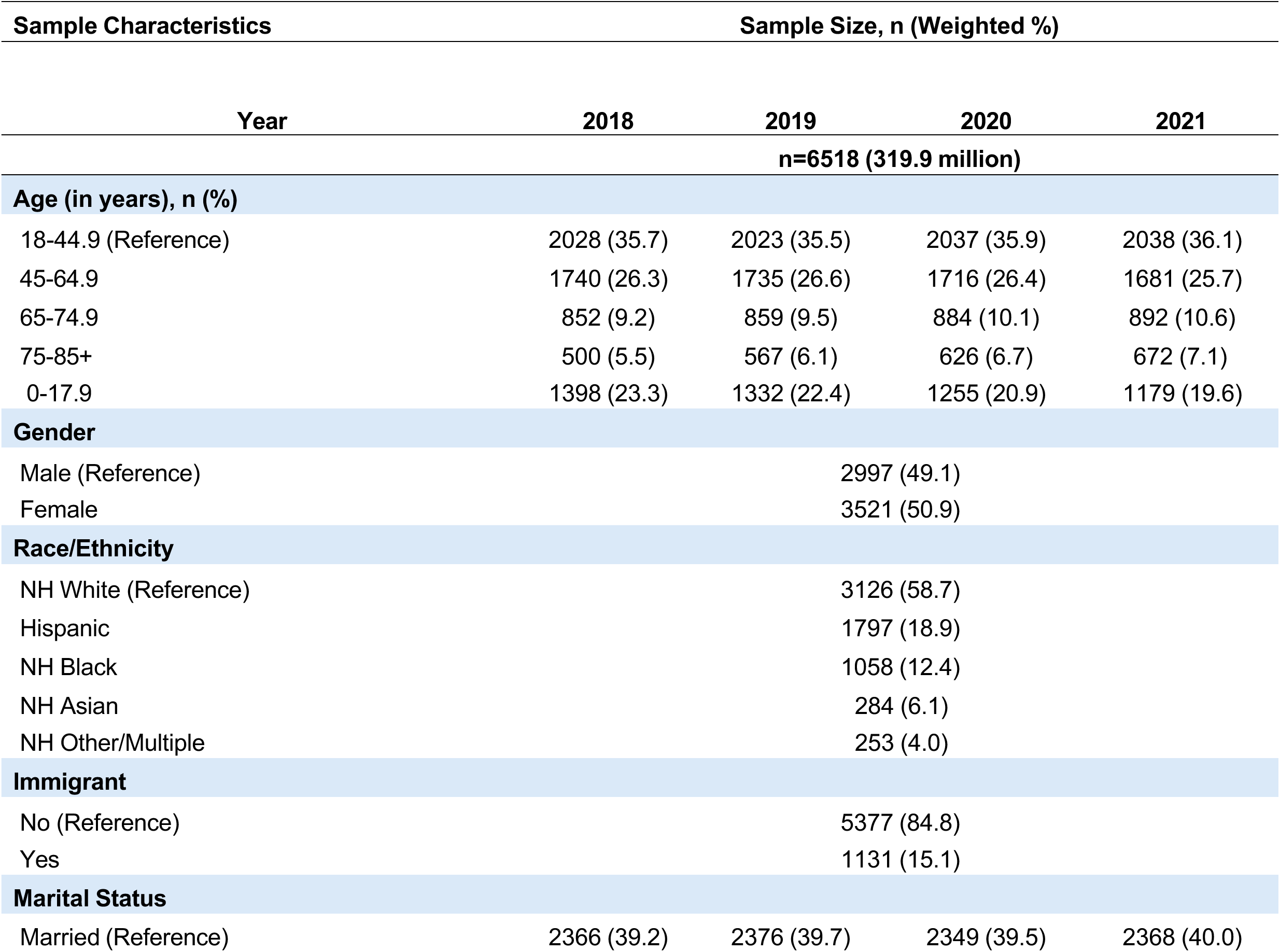
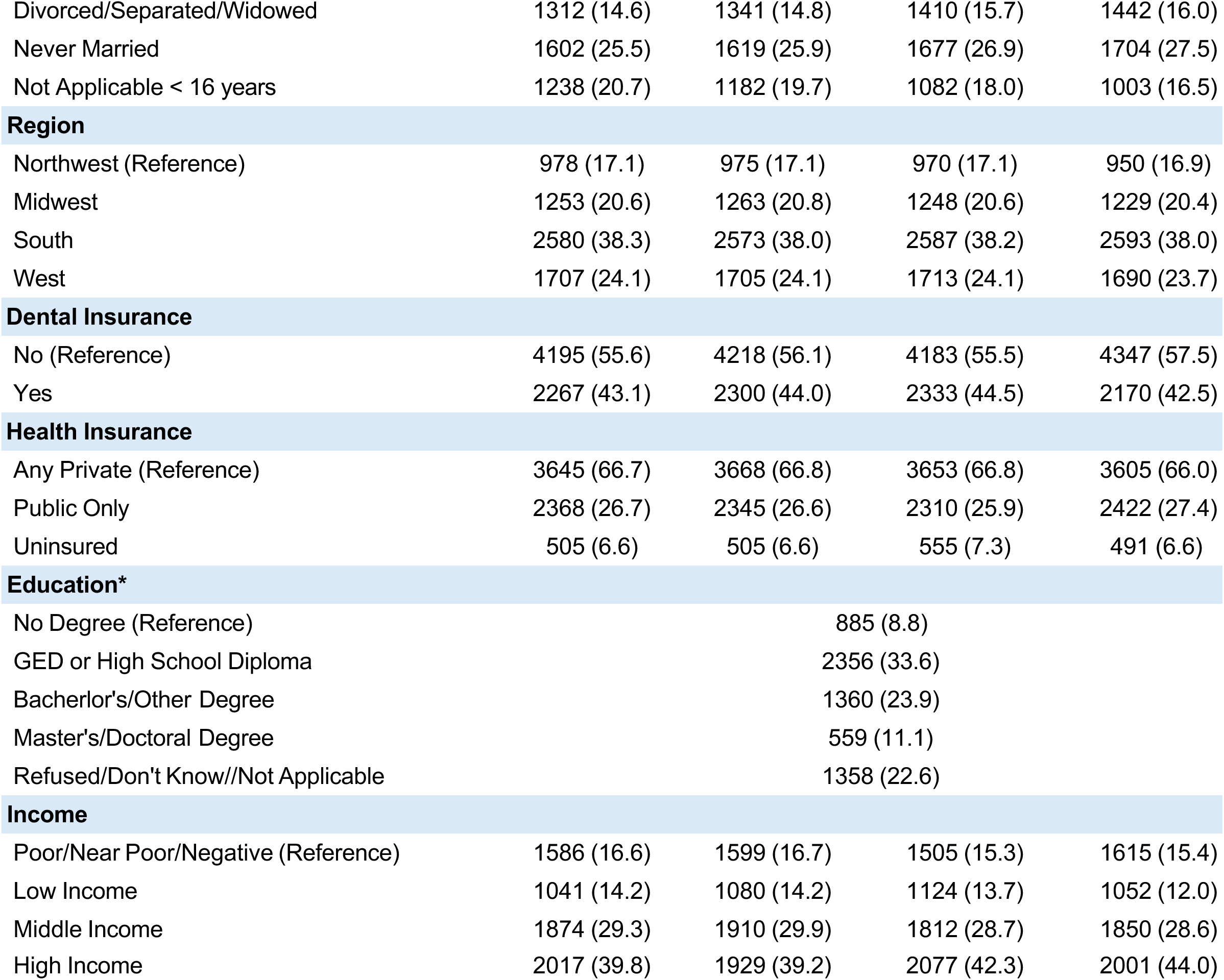
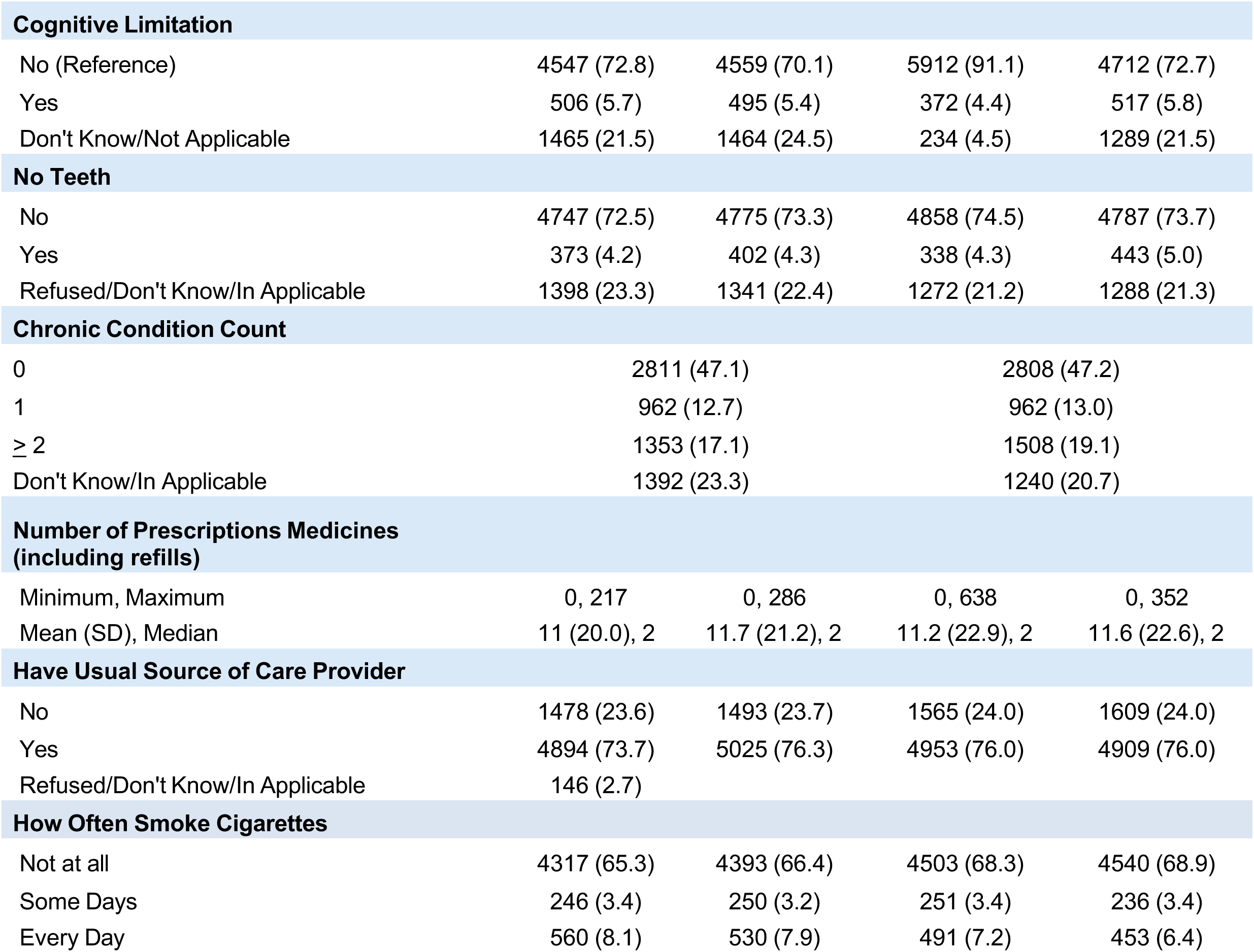
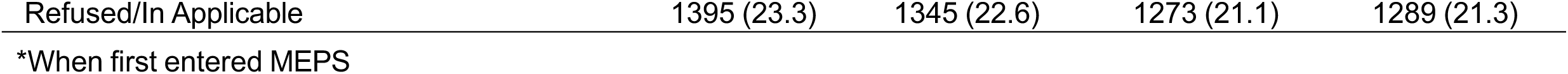
Characteristics of the U.S. Population, Medical Expenditure Panel Survey, Years 2018-2021.

| Sample Characteristics |  | Sample Size, n (Weighted %) |  |  |
| --- | --- | --- | --- | --- |
| Year | 2018 | 2019 | 2020 | 2021 |
| n=6518 (319.9 million) |  |  |  |  |
| Age (in years), n (%) |  |  |  |  |
| 18-44.9 (Reference) | 2028 (35.7) | 2023 (35.5) | 2037 (35.9) | 2038 (36.1) |
| 45-64.9 | 1740 (26.3) | 1735 (26.6) | 1716 (26.4) | 1681 (25.7) |
| 65-74.9 | 852 (9.2) | 859 (9.5) | 884 (10.1) | 892 (10.6) |
| 75-85+ | 500 (5.5) | 567 (6.1) | 626 (6.7) | 672 (7.1) |
| 0-17.9 | 1398 (23.3) | 1332 (22.4) | 1255 (20.9) | 1179 (19.6) |
| Gender |  |  |  |  |
| Male (Reference) |  | 2997 (49.1) |  |  |
| Female |  | 3521 (50.9) |  |  |
| Race/Ethnicity |  |  |  |  |
| NH White (Reference) |  | 3126 (58.7) |  |  |
| Hispanic |  | 1797 (18.9) |  |  |
| NH Black |  | 1058 (12.4) |  |  |
| NH Asian |  | 284 (6.1) |  |  |
| NH Other/Multiple |  | 253 (4.0) |  |  |
| Immigrant |  |  |  |  |
| No (Reference) |  | 5377 (84.8) |  |  |
| Yes |  | 1131 (15.1) |  |  |
| Marital Status |  |  |  |  |
| Married (Reference) | 2366 (39.2) | 2376 (39.7) | 2349 (39.5) | 2368 (40.0) |
| Divorced/Separated/Widowed | 1312 (14.6) | 1341 (14.8) | 1410 (15.7) | 1442 (16.0) |
| Never Married | 1602 (25.5) | 1619 (25.9) | 1677 (26.9) | 1704 (27.5) |
| Not Applicable < 16 years | 1238 (20.7) | 1182 (19.7) | 1082 (18.0) | 1003 (16.5) |
| **Region** |  |  |  |  |
| Northwest (Reference) | 978 (17.1) | 975 (17.1) | 970 (17.1) | 950 (16.9) |
| Midwest | 1253 (20.6) | 1263 (20.8) | 1248 (20.6) | 1229 (20.4) |
| South | 2580 (38.3) | 2573 (38.0) | 2587 (38.2) | 2593 (38.0) |
| West | 1707 (24.1) | 1705 (24.1) | 1713 (24.1) | 1690 (23.7) |
| **Dental Insurance** |  |  |  |  |
| No (Reference) | 4195 (55.6) | 4218 (56.1) | 4183 (55.5) | 4347 (57.5) |
| Yes | 2267 (43.1) | 2300 (44.0) | 2333 (44.5) | 2170 (42.5) |
| **Health Insurance** |  |  |  |  |
| Any Private (Reference) | 3645 (66.7) | 3668 (66.8) | 3653 (66.8) | 3605 (66.0) |
| Public Only | 2368 (26.7) | 2345 (26.6) | 2310 (25.9) | 2422 (27.4) |
| Uninsured | 505 (6.6) | 505 (6.6) | 555 (7.3) | 491 (6.6) |
| **Education\*** |  |  |  |  |
| No Degree (Reference) |  | 885 (8.8) |  |  |
| GED or High School Diploma |  | 2356 (33.6) |  |  |
| Bachelor's/Other Degree |  | 1360 (23.9) |  |  |
| Master's/Doctoral Degree |  | 559 (11.1) |  |  |
| Refused/Don't Know//Not Applicable |  | 1358 (22.6) |  |  |
| **Income** |  |  |  |  |
| Poor/Near Poor/Negative (Reference) | 1586 (16.6) | 1599 (16.7) | 1505 (15.3) | 1615 (15.4) |
| Low Income | 1041 (14.2) | 1080 (14.2) | 1124 (13.7) | 1052 (12.0) |
| Middle Income | 1874 (29.3) | 1910 (29.9) | 1812 (28.7) | 1850 (28.6) |
| High Income | 2017 (39.8) | 1929 (39.2) | 2077 (42.3) | 2001 (44.0) |
| **Cognitive Limitation** |  |  |  |  |
| No (Reference) | 4547 (72.8) | 4559 (70.1) | 5912 (91.1) | 4712 (72.7) |
| Yes | 506 (5.7) | 495 (5.4) | 372 (4.4) | 517 (5.8) |
| Don't Know/Not Applicable | 1465 (21.5) | 1464 (24.5) | 234 (4.5) | 1289 (21.5) |
| **No Teeth** |  |  |  |  |
| No | 4747 (72.5) | 4775 (73.3) | 4858 (74.5) | 4787 (73.7) |
| Yes | 373 (4.2) | 402 (4.3) | 338 (4.3) | 443 (5.0) |
| Refused/Don't Know/In Applicable | 1398 (23.3) | 1341 (22.4) | 1272 (21.2) | 1288 (21.3) |
| **Chronic Condition Count** |  |  |  |  |
| 0 | 2811 (47.1) |  | 2808 (47.2) |  |
| 1 | 962 (12.7) |  | 962 (13.0) |  |
| ≥ 2 | 1353 (17.1) |  | 1508 (19.1) |  |
| Don't Know/In Applicable | 1392 (23.3) |  | 1240 (20.7) |  |
| **Number of Prescriptions Medicines (including refills)** |  |  |  |  |
| Minimum, Maximum | 0, 217 | 0, 286 | 0, 638 | 0, 352 |
| Mean (SD), Median | 11 (20.0), 2 | 11.7 (21.2), 2 | 11.2 (22.9), 2 | 11.6 (22.6), 2 |
| **Have Usual Source of Care Provider** |  |  |  |  |
| No | 1478 (23.6) | 1493 (23.7) | 1565 (24.0) | 1609 (24.0) |
| Yes | 4894 (73.7) | 5025 (76.3) | 4953 (76.0) | 4909 (76.0) |
| Refused/Don't Know/In Applicable | 146 (2.7) |  |  |  |
| **How Often Smoke Cigarettes** |  |  |  |  |
| Not at all | 4317 (65.3) | 4393 (66.4) | 4503 (68.3) | 4540 (68.9) |
| Some Days | 246 (3.4) | 250 (3.2) | 251 (3.4) | 236 (3.4) |
| Every Day | 560 (8.1) | 530 (7.9) | 491 (7.2) | 453 (6.4) |
| Refused/In Applicable | 1395 (23.3) | 1345 (22.6) | 1273 (21.1) | 1289 (21.3) |

\*When first entered MEPS

### 3.1 Age-Related Trends in Dental Visits

From 2018 to 2021, annual dental visit rates among adults rose slightly in every adult age group: from 15% to 16% for ages 18-44, from 5% to 6% for ages 65-74.9, and from 3% to 4% for those 75 and older (p < 0.0001). By contrast, the percentage of children (0-17) with at least one dental visit declined from 13% to 11%. For those who accessed dental care, the average number of visits per year generally decreased across most groups, including children (2.2 to 2.1), middle-aged adults (2.2 to 2.1), and older adults aged 65–74 (2.6 to 2.5), but rose modestly among younger adults (1.9 to 2.0). (**Figure 2**, **Table 3**)

**Figure 2:**
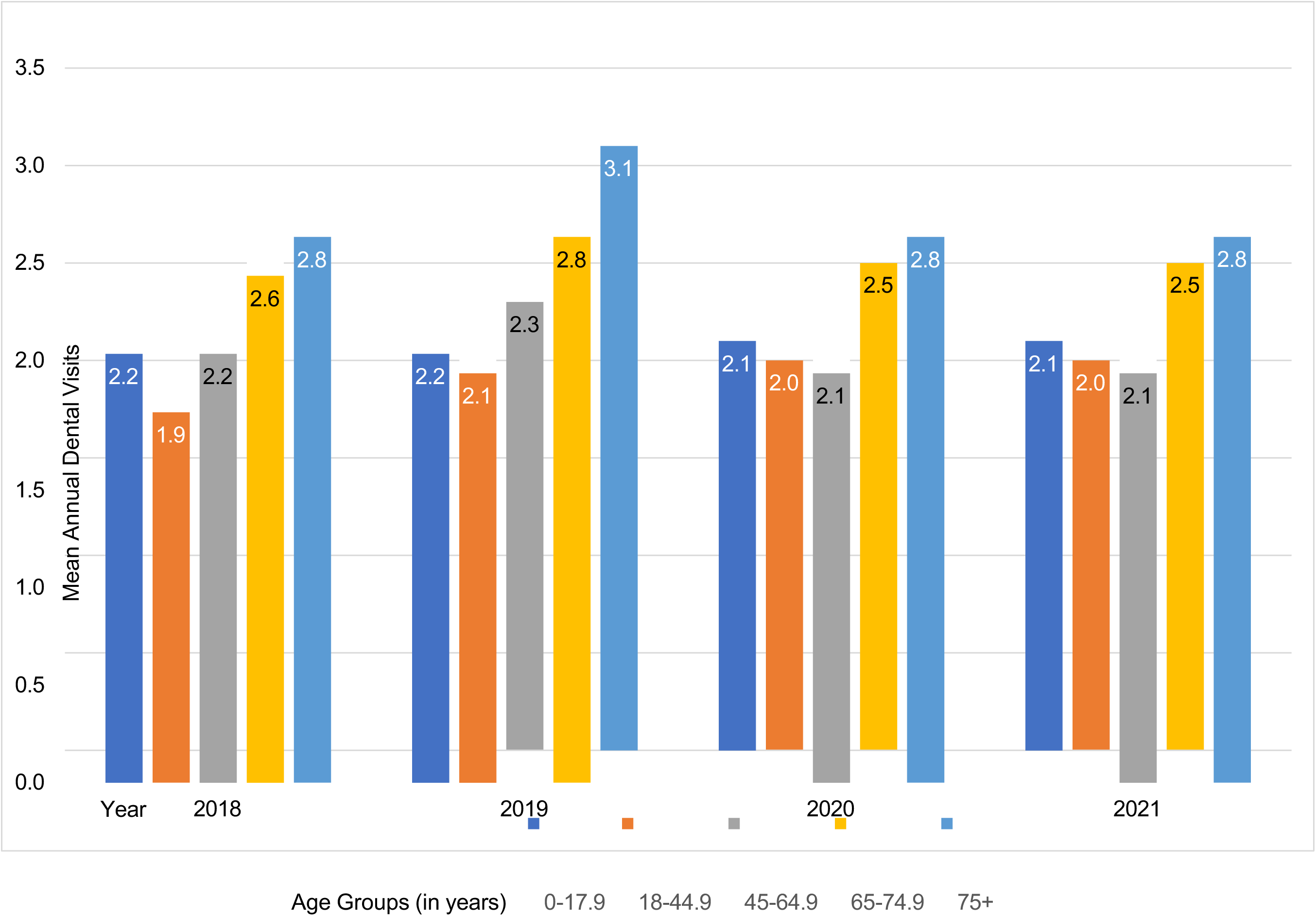
Survey Weighted Mean Annual Dental Visits Among Those Who Had At least One Dental Visit by Age Group and Year, Medical Expenditure Panel Survey, Years 2018-2021.

**Table 3:** Survey Weighted Mean Dental Visits Among those who had at least one Dental Visit by Age Group and Year, Medical Expenditure Panel Survey, Years 2018-2021.

| Year | n<br>(Weighted %) | Dental<br>Visits $\geq 1$ | Age Groups (in years) | | | | |
| --- | --- | --- | --- | --- | --- | --- | --- |
|  |  |  | 0-17.9 | 18-44.9 | 45-64.9 | 65-74.9 | 75+ |
| 2018 | 2961 (48.9) | Mean<br>(95% CI) | 2.2 (2.0, 2.4) | 1.9 (1.8, 2.0) | 2.2 (2.0, 2.3) | 2.6 (2.4, 2.8) | 2.8 (2.4, 3.3) |
| 2019 | 2837 (47.0) |  | 2.2 (2.0, 2.4) | 2.1 (1.9, 2.2) | 2.3 (2.1, 2.4) | 2.8 (2.6, 3.7) | 3.1 (2.8, 3.5) |
| 2020 | 2961 (48.9) |  | 2.1 (1.9, 2.4) | 2.0 (1.9, 2.1) | 2.1 (2.0, 2.3) | 2.5 (2.3, 2.7) | 2.8 (2.4, 3.2) |
| 2021 | 2961 (48.9) |  | 2.1 (1.7, 3.3) | 2.0 (1.9, 2.1) | 2.1 (2.0, 2.3) | 2.5 (2.3, 2.7) | 2.8 (2.4, 3.1) |

### 3.2 Poisson Regression Analysis

The association between age group and dental visits was consistently stronger in the unadjusted Poisson regression models than in the adjusted models (**Tables 4-5**). When confounding variables were controlled for, the prevalence ratio (PR) for dental visits declined from 2018 to 2021 among adults and older adults, but there was a pronounced increase for children in 2021 (PR 8.41; 95% CI, 4.0-17.9). Among adults aged 65-74.9, the PR rose from 1.57 in 2018 to 1.91 in 2020, then returned toward pre-pandemic levels in 2021 (PR 1.63; 95% CI, 1.3-2.0). Adults aged 75 and older showed a similar trend, with the PR peaking at 2.13 in 2020 and decreasing to 1.66 in 2021. Further details on these trends, including percentage changes in dental care use, are presented in **Table 6**.

**Table 4:** Unadjusted Poisson Regression Analysis of Dental Care Visits, Medical Expenditure Panel Survey, Years 2018-2021.

| Sample Size, n (Survey Weighted) |  | 6518 (319.9 million) |  |  |
| --- | --- | --- | --- | --- |
| Year | 2018 | 2019 | 2020 | 2021 |
| Total Dental Visits | Prevalence Ratio (95% CI) |  |  |  |
| Age (in years), n (Weighted %) |  |  |  |  |
| 18-44.9 (Ref) |  |  |  |  |
| 0-17.9 | 1.48 (1.3, 1.7) | 1.49 (1.3, 1.7) | 1.74 (1.4, 2.1) | 1.84 (1.6, 2.1) |
| 45-64.9 | 1.25 (1.1, 1.4) | 1.21 (1.1, 1.4) | 1.36 (1.1, 1.6) | 1.35 (1.2, 1.6) |
| 65-74.9 | 1.84 (1.6, 2.1) | 1.98 (1.7, 2.3) | 2.04 (1.7, 2.5) | 1.94 (1.7, 2.3) |
| 75-85+ | 1.86 (1.6, 2.1) | 1.94 (1.6, 2.3) | 2.01 (1.6, 2.5) | 1.88 (1.5, 2.3) |

\*Bold indicates statistical significance, $p < 0.001$
2018, F (4, 137), Prob>F, <0.001
2019, F( 5, 136), Prob>F, <0.001
2020, F (4, 137), Prob>F, <0.001
2021, F(5, 136 ), Prob>F, <0.001

**Table 5:** Adjusted Poisson regression analysis of dental care visits pre- and during COVID-19, Medical Expenditure Panel Survey, Years 2018-2021.

| Sample Size, n (Survey Weighted) | 6518 (319.9 million) |  |  |  |
| --- | --- | --- | --- | --- |
| Year | 2018 | 2019 | 2020 | 2021 |
| Total Dental Visits | Prevalence Ratio (95% CI) |  |  |  |
| Age (in years), n (Weighted %) |  |  |  |  |
| 18-44.9 (Ref) |  |  |  |  |
| 0-17.9 | 1.64 (0.7, 3.7) | 10.18 (0.7, 141) | 1.80 (0.6, 5.1) | **8.41 (4.0, 17.9)** |
| 45-64.9 | 1.08 (0.9, 1.2) | 1.04 (0.9, 1.2) | 1.22 (1.0, 1.5) | 1.11 (0.9, 1.3) |
| 65-74.9 | **1.57 (1.3, 1.9)** | **1.72 (1.4, 2.1)** | **1.91 (1.5, 2.4)** | **1.63 (1.3, 2.0)** |
| 75-85+ | **1.73 (1.4, 2.1)** | **1.84 (1.5, 2.3)** | **2.13 (1.7, 2.7)** | **1.66 (1.3, 2.0)** |
| Gender |  |  |  |  |
| Male (Ref) |  |  |  |  |
| Female | **1.16 (1.1, 1.3)** | 1.12 (1.0, 1.2) | 1.13 (1.0, 1.3) | 1.12 (1.0, 1.3) |
| Race/Ethnicity |  |  |  |  |
| NH White (Ref) |  |  |  |  |
| Hispanic | 0.79 (0.7, 0.9) | 0.80 (0.7, 1.0) | **0.65 (0.5, 0.8)** | **0.61 (0.5, 0.7)** |
| NH Black | **0.71 (0.6, 0.9)** | 0.77 (0.6, 1.0) | **0.67 (0.5, 0.9)** | **0.70 (0.6, 0.9)** |
| NH Asian | **0.65 (0.5, 0.9)** | 0.75 (0.5, 1.1) | **0.55 (0.4, 0.8)** | **0.52 (0.4, 0.7)** |
| NH Other/Multiple | **0.86 (0.6, 1.2)** | 0.91 (0.6, 1.3) | 0.65 (0.4, 1.0) | 0.72 (0.5, 1.0) |
| Immigrant |  |  |  |  |
| No (Ref) |  |  |  |  |
| Yes | 0.90 (0.7, 1.1) | 0.97 (1.0, 1.2) | 0.84 (0.7, 1.0) | 1.04 (0.8, 1.3) |
| Marital Status |  |  |  |  |
| Married (Ref) |  |  |  |  |
| Divorced/Separated/Widowed | 0.99 (0.9, 1.1) | 1.02 (0.9, 1.2) | 1.10 (0.9, 1.3) | 1.06 (0.9, 1.3) |
| Never Married | 1.08 (0.9, 1.2) | 0.91 (0.8, 1.1) | 1.08 (0.9, 1.2) | 1.01 (0.8, 1.2) |
| Region |  |  |  |  |
| Northwest (Ref) |  |  |  |  |
| Midwest | 1.21 (1.0, 1.5) | 0.96 (0.8, 1.2) | 1.22 (1.0, 1.5) | 1.01 (0.8, 1.3) |
| South | 0.92 (0.8, 1.1) | 0.81 (0.7, 1.0) | 0.94 (0.8, 1.2) | 0.80 (0.7, 1.0) |
| West | 1.18 (1.0, 1.4) | 0.97 (0.8, 1.2) | 1.28 (1.0, 1.6) | 1.04 (0.8, 1.3) |
| Dental Insurance |  |  |  |  |
| No (Ref) |  |  |  |  |
| Yes | 1.10 (1.0, 1.3) | 1.08 (0.8, 1.2) | **1.28 (1.1, 1.5)** | **1.19 (1.0, 1.4)** |
| Health Insurance |  |  |  |  |
| Any Private (Reference) |  |  |  |  |
| Public Only | 0.90 (0.8, 1.1) | 0.90 (0.8, 1.0) | 0.87 (0.7, 1.0) | 0.89 (0.7, 1.0) |
| Uninsured | 0.85 (0.6, 1.3) | 0.91 (0.6, 1.4) | 0.88 (0.6, 1.3) | 0.52 (0.4, 0.8) |
| Education* |  |  |  |  |
| No Degree (Ref) |  |  |  |  |
| GED or High School Diploma | **1.41 (1.1, 1.8)** | **1.63 (1.2, 2.1)** | 1.0 (0.8, 1.4) | 1.34 (1.0, 1.8) |
| Bachelor's/Other Degree | **1.68 (1.3, 2.2)** | **1.98 (1.5, 2.6)** | 1.40 (1.1, 1.9) | **1.44 (1.1, 1.9)** |
| Master's/Doctoral Degree | **1.73 (1.3, 2.3)** | **2.26 (1.7, 3.0)** | 1.47 (1.1, 2.0) | **1.68 (1.2, 2.3)** |
| Income |  |  |  |  |
| Poor/Near Poor/Negative (Ref) |  |  |  |  |
| Low Income | 0.96 (0.8, 1.2) | 1.10 (0.9, 1.4) | 1.01 (0.8, 1.3) | 1.0 (0.8, 1.3) |
| Middle Income | 1.26 (1.0, 1.6) | **1.45 (1.2, 1.8)** | 1.19 (0.9, 1.5) | **1.0 (1.1, 1.7)** |
| High Income | **1.44 (1.2, 1.8)** | **1.68 (1.4, 2.0)** | **1.51 (1.2, 1.9)** | **1.63 (1.3, 2.1)** |
| Cognitive Limitation |  |  |  |  |
| No (Ref) |  |  |  |  |
| Yes | 1.13 (0.9, 1.4) | 0.92 (0.7, 1.1) | 0.97 (0.7, 1.2) | 0.8 (0.6, 1.0) |
| No Teeth |  |  |  |  |
| No |  |  |  |  |
| Yes | **0.42 (0.3, 0.6)** | **0.35 (0.2, 0.5)** | **0.38 (0.2, 0.6)** | **0.57 (0.4, 0.9)** |
| Chronic Condition Count |  |  |  |  |
| 0 (Ref) |  |  |  |  |
| 1 | 1.06 (0.9, 1.2) | 0.93 (0.8, 1.1) | 0.96 (0.8, 1.1) | 0.84 (0.7, 1.0) |
| ≥ 2 | 1.00 (0.9, 1.2) | 0.89 (0.8, 1.0) | 0.90 (0.8, 1.1) | 1.03 (0.9, 1.2) |
| Number of Prescriptions Medicines (including refills) |  |  |  |  |
| Minimum, Maximum | 1.00 (1.0, 1.0) | **1.01 (1.0, 1.0)** | **1.00 (1.0, 1.0)** | **1.00 (1.0, 1.0)** |
| Mean (SD), Median |  |  |  |  |
| Have Usual Source of Care Provider |  |  |  |  |
| No (Ref) |  |  |  |  |
| Yes | **1.46 (1.3, 1.7)** | **1.28 (1.1, 1.5)** | **1.35 (1.1, 1.7)** | **1.36 (1.1, 1.6)** |
| How Often Smoke Cigarettes |  |  |  |  |
| Not at all (Ref) |  |  |  |  |
| Some Days | 0.68 (0.5, 1.0) | 0.91 (0.6, 1.4) | 0.81 (0.5, 1.3) | 0.76 (0.5, 1.2) |
| Every Day | 0.84 (0.7, 1.0) | **0.71 (0.6, 0.9)** | **0.57 (0.4, 0.8)** | **0.68 (0.5, 0.9)** |

\*When first entered MEPS
Bold indicates statistical significance, $p \leq 0.001$
2018, F (41, 100), $\text{Prob} > F, \leq 0.01$
2019, F (40, 101), $\text{Prob} > F, \leq 0.01$
2020, F (40, 101), $\text{Prob} > F, \leq 0.01$
2021, F (41, 100), $\text{Prob} > F, \leq 0.01$

**Table 6:**
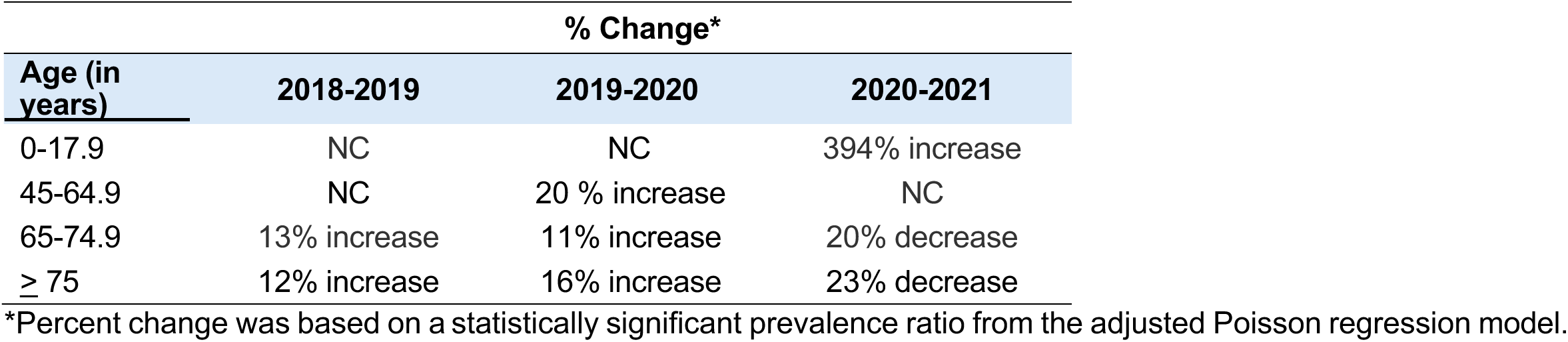
Percent change in survey-weighted mean annual dental visits, Medical Expenditure Panel Survey, Years 2018-2021.

### 3.3 Factors Associated with Dental Care Visits

Several factors were linked to changes in dental care use during the COVID-19 period. Increased dental visits were associated with having dental insurance, higher income, and a usual source of care. In contrast, lower utilization was observed among individuals who smoked daily, had lost all teeth, or self-identified as Hispanic, Black, or Asian. Educational attainment did not significantly impact dental care use during the pandemic peak.

## 4. Discussion

This analysis, based on four years of nationally representative survey data, provides population-level estimates of dental care utilization across five age cohorts in the U.S. after adjusting for multiple confounding variables. Findings partially supported the initial hypothesis: average annual dental visits decreased in all age groups from 2019 to 2020, but only partially recovered in 2021. The adjusted Poisson regression revealed increased dental care visits among adults aged 65 and older in 2020 and among children in 2021. The uptick in visits by older adults in 2020 may signal a greater need for urgent or emergency dental procedures, possibly due to delays from earlier lockdowns. By 2021, dental care use among those aged 65-74 and 75+ rebounded less rapidly than among children, possibly reflecting continued COVID-19 concerns among older populations. These trends highlight the need for ongoing monitoring of dental care use after 2021 and assessment of the long-term health effects of deferred dental care. The rise in children’s dental visits in 2021 may be attributed to increased parental confidence in seeking preventive care as clinics reopened and vaccines became available. Furthermore, the backlog of dental problems such as gum disease and cavities, resulting from postponed care, might have fueled additional visits in 2021.

### 4.1 Similar Studies

Numerous prior studies have analyzed national survey data to examine dental care utilization in the U.S. population. However, only a limited number have assessed these patterns across comparable age cohorts while accounting for a comprehensive array of factors throughout the COVID-19 pandemic [28,29,42,43]. For example, one descriptive MEPS report (2019-2021) found that dental visits per person declined across all age groups in 2020 and rebounded in 2021 [28]. That report also noted that among those who accessed dental care, individuals aged 75 and older had the highest average number of visits, followed by those aged 65-74 [28]. Longitudinal research has shown that regular dental visits can mitigate racial and socioeconomic disparities in tooth loss, highlighting the public health value of consistent dental care [45]. In line with these findings, our results show a similar dip in average annual dental visits in 2020 among dental care users across all age groups, with the highest utilization rates observed in older adults.

An ADA report using MEPS data demonstrated that dental visits declined among children, adults, and older adults in 2020 but rebounded in both 2021 and 2022, with the highest rates consistently observed among children and older adults compared with adults across all years [29]. That report estimated that 45% of Americans visited a dentist in 2022, a slight increase from 2021. In our study, the 2021 estimate was higher, with about 49% of the U.S. population, roughly 163 million people, having a dental care visit. Another investigation found that the likelihood of dental visits among middle-aged and older adults declined by nearly 5% from 2019 to 2020, highlighting the need for further research on pandemic-related trends [42]. Machine learning research further reveals persistent gender and racial disparities in dental services use, emphasizing the need for fairness-oriented strategies in dental care [46]. Consistent with these prior findings, we observed that older adults had consistently lower proportions of dental care visits compared to children and adults, yet our adjusted Poisson regression indicated a dose-dependent rise in dental care use among adults aged 65–74 and 75+ during 2018–2020, with those 75 and older maintaining the highest utilization. Additional evidence suggests that within-person dental visits decreased overall and across various sociodemographic and economic groups between 2019 and 2020 [44]. In our analysis, higher education was associated with increased dental care visits before COVID-19 (2018–2019) and in the second pandemic year (2021), but not in 2020. Only individuals with high incomes were likely to obtain dental care in 2020, possibly for urgent or non-preventive reasons, and only if they could afford treatment. Dental insurance and a usual source of care were associated with greater dental care use across all years, underscoring the importance of insurance coverage for access to oral health services.

### 4.2 Clinical Significance

Clinical significance: Reducing barriers to dental care utilization across all age groups is essential for restorative dental practice and oral health policy. Our findings demonstrate that the COVID-19 pandemic led to significant disruptions in dental service use, particularly among older adults, with only partial recovery by 2021. This highlights the need for clinician- and policy-driven strategies that address access, especially for preventive and restorative services, as well as risk assessment and prevention strategies, to minimize long-term impacts on caries burden, untreated decay, and tooth loss. Future research should focus on identifying modifiable risk and protective clinical strategies in consultation with dentists that influence delayed care and oral disease management, with an emphasis on adults and older adults.

### 4.3 Strengths and Limitations

A major strength of this study is the use of a nationally representative, longitudinal MEPS Panel 23 cohort spanning four years, which enables a comprehensive assessment of COVID-19’s impact on dental care patterns in the U.S. population. The survey’s weighting procedures enhance the generalizability of the results to national trends. The American Dental Association’s Health Policy Institute also relies on MEPS, noting that alternative data sources such as NHIS, NHANES, and BRFSS may be less reliable and could overestimate dental care use [29]. Several limitations are noteworthy. First, while MEPS includes a household verification step, most data are self-reported and thus subject to recall bias, especially during the stress of the pandemic. Second, although survey weights address non-response, reductions in MEPS participation during COVID-19 [35] may have introduced potential bias or measurement error, possibly underestimating dental care use. Third, our reliance on annual data means early 2020 (pre-pandemic) months are included, potentially understating the effect of the initial lockdown. Fourth, our year-by-year analysis cannot account for within-person variation and may overlook individual-level changes. Fifth, MEPS provides only binary gender classification. Additionally, qualitative research indicates that both professional and structural barriers can hinder the implementation of preventive dental care in practice, suggesting the need for system-level reforms to ensure equitable service provision [47]. Lastly, while we controlled for many confounders, the analysis is observational and cannot establish causality.

### 4.4 Conclusion

This study provides nationally representative evidence that dental care visits in the U.S. declined during the COVID-19 pandemic, with incomplete rebound by 2021 and marked disparities persisting among older adults. These patterns have important implications for restorative dentistry, highlighting the need to integrate risk assessment and prevention strategies to reduce the risk of caries progression, tooth loss, and unmet prosthodontic needs in older populations. Strategies to improve access and continuity of dental services, informed by ongoing research into factors that influence dental use, are critical for both clinical practice and oral health policy as the profession moves beyond the pandemic.

## Data Availability

All data produced are available online at https://meps.ahrq.gov/mepsweb/

## 5. Declaration of generative AI and AI-assisted technologies in the manuscript preparation process

No generative AI tools were used for study design, data curation, or analysis, creating any figures, tables, or writing the original draft. Google Gemini 3.5 Flash was used to revise the introduction and to ensure coherence with prior research. The corresponding author reviewed all content after editing and takes full responsibility for the article’s originality and integrity.

## Acknowledgments

We thank the 2025 AcademyHealth Annual Research Meeting, the 2022-2025 Agency for Healthcare Research and Quality (AHRQ) Medical Expenditure Panel Survey (MEPS) workshop speakers, and the feedback received at the Collaborative for Innovation in Data & Measurement in Aging (CIDMA) workshops from 2022-2025. We thank Sharon Larson, PhD, David J. Whellan, MD, MHS, FACC, FAHA, Jeanne M. Felter, PhD, LPC, Billy Oglesby, PhD, MBA, MSPH, FACHE for providing PPZ Principal Investigator status and the support to carry out the proposed research from NIA as a sub-award and as a Visiting Researcher at Thomas Jefferson University. Special thanks to Charlene Ruffin and Holly Talley at Thomas Jefferson University, Philadelphia, PA, for their help with pre-award and post-award submissions. This manuscript is based on an earlier version of this study, which was published as a preprint and has not undergone formal peer review [48].

## CRediT Authorship Contribution Statement

Preeti Pushpalata Zanwar: Conceptualization, Methodology, Validation, Formal Analysis,

Investigation, Resources, Data Curation, Writing – Original Draft, Writing-Review & Editing,

Visualization, Supervision, Project Administration, Funding Acquisition

Hossain Zare: Methodology, Writing – Review & Editing, Supervision

Kopal Mathur: Writing – Review & Editing

Lyubov Slashcheva: Writing – Review & Editing

Bei Wu: Writing –Review & Editing, Supervision

## Data statement

The data files used in the paper are publicly available from the Medical Expenditure Panel Survey https://meps.ahrq.gov/mepsweb/

